# Per- and poly-fluoroalkyl substances and bone mineral density: results from the Bayesian weighted quantile sum regression

**DOI:** 10.1101/19010710

**Authors:** Elena Colicino, Nicolo Foppa Pedretti, Stefanie Busgang, Chris Gennings

## Abstract

**Background:** Per- and poly-fluoroalkyl substances (PFAS) are chemicals, detected in 95% of Americans, that induce osteotoxicity and modulate hormones thereby influencing bone health. Previous studies found associations between individual PFAS and bone mineral density but did not analyze their combined effects.

**Objective:** To extend weighted quantile sum (WQS) regression to a Bayesian framework (BWQS) and determine the association between a mixture of serum PFAS and mineral density in lumbar spine, total and neck femur in 499 adults from the 2013–2014 National Health and Nutrition Examination Survey (NHANES).

**Methods:** We used BWQS to assess the combined association of nine PFAS, as a mixture, with bone mineral density in adults. As secondary analyses, we focused on vulnerable populations (men over 50 years and postmenopausal women). Analyses were weighted according to NHANES weights and were adjusted for socio-demographic factors. Sensitivity analyses included bone mineral density associations with individual compounds and results from WQS regressions.

**Results:** The mean age was 55 years old (Standard Error [SE]=1) with average spine, total and neck femur mineral densities of 1.01 (SE=0.01), 0.95 (SE=0.01), and 0.78 (SE=0.01) gm/cm^2^, respectively. PFAS mixture levels showed no evidence of association with mineral density (spine: β=-0.004; 95% credible interval [CrI]=-0.04, 0.04; total femur: β=0.002; 95%CrI=-0.04, 0.05; femur neck: β=0.005; 95%CrI=-0.03, 0.04) in the overall population. Results were also null in vulnerable populations. Findings were consistent across sensitivity analyses.

**Conclusions:** We introduced a Bayesian extension of WQS and found no evidence of the association between PFAS mixture and bone mineral density.

## Introduction

As the population ages, low bone mineral density has emerged as a public health concern because it is related to fractures, morbidities, hospitalizations, and premature mortality^.1,2^ Deterioration of bone mass differs between the sexes, with a higher prevalence in women. About 10% of women over 50 years of age suffer from low bone mineral density, but only 2% of men of the same age have similar bone deterioration.^3^ Traditional risk factors associated with decreased bone mineral density include chronological age, family history of bone disease, suboptimal high-impact physical activity, and smoking.^4^ However, recent evidence suggests that environmental exposures, including air pollution, lead, cadmium, and mercury, are associated with lower bone mineral density and higher risks for osteoporosis.^5-7^

Per- and poly-fluoroalkyl substances (PFAS) are synthetic organic chemicals that have been used extensively in industrial processes and commercial applications since the 1950s. These chemicals are widely used and persist for long periods of time, resulting in increased levels of environmental contamination.^8,9^ The primary sources of human PFAS exposure include migration from food packaging and cookware, drinking water, indoor air, and house dust.^10^ PFAS have been detected *in vivo* in human tissue samples,^11,12^ in 95% of the U.S. population.^13^ PFAS are poorly metabolized and excreted slowly from the human body, with half-lives of 4–8 years.^14^ Previous human and animal studies showed that PFAS bioaccumulate in bones, with perfluorooctanoate (PFOA) being predominant.^15,16^ Due to their limited susceptibility to degradation and slow elimination by human bodies, human exposure to PFAS is of increasing concern.^17,18^

The toxicity of PFAS to bones has been reported in human and animal studies, with high PFAS concentrations associated with adverse skeletal outcomes, suggesting that bones are target tissues for PFAS toxicity.^19^ PFAS are also endocrine-disrupting chemicals ^20^ based on their hormonal modulation. ^21^ PFAS have been suggested to impact bone density via sex hormones.^22,23^ In rodents, exposure to PFAS was negatively associated with bone structure and biomechanical properties.^24^ Bone cell cultures showed increased bone resorption activity at low, albeit environmentally relevant, PFAS concentrations in human bone marrow and peripheral blood–derived osteoclasts, through the effect of PFAS on the cytokine and clastokine profiles during cell differentiation.^19^ In human studies, bone PFAS concentrations and relative bone volume have been correlated, but results are still inconclusive. In a few cross-sectional studies, serum levels of a few PFAS were negatively associated with bone mineral density only in women,^19,21^ but findings for most compounds are null in the general U.S. population.^19,21^ In addition, all previous studies focused on the association between individual PFAS and bone health outcomes; however, given their ubiquity and persistence, exposure likely occurs to many PFAS simultaneously. Those studies failed to account for the correlation structure among PFAS or to consider PFAS exposure as a mixture.

Environmental health studies have applied weighted quantile sum (WQS) regression to assess the mixture effect of multiple co-occurring factors and to identify the driving factors in the mixture. Briefly, the WQS regression summarizes the overall exposure to the mixture by estimating a single weighted index and accounts for the individual contribution of each component of the mixture by using weights.^25,26^ WQS regression splits the dataset into two subsets, a training set (generally 40%) and a validation set (60%). In the first set, this approach estimates the weights using an ensemble step and estimates average weights across B bootstrap samples, and in the second subset it estimates the coefficient mapped to the mixture, conditionally to the estimated weights.^25-27^ This regression also requires a priori selection of the directionality (positive or negative) of the coefficient associated with the mixture, and it conditions on the weights in the weighted index for testing for significance using the hold out validation set. As such, it does not provide diagnostics (confidence intervals, standard deviation, and p values) about the weights of the components of the mixture.^25,26^

Here we proposed a novel Bayesian extension of the WQS regression (BWQS) to overcome its limitations and to illustrate the method for assessing the combined association of several forms of serum PFAS as a mixture with bone mineral density in 499 U.S. adults from the 2013–2014 cycle of the National Health and Nutrition Examination Survey (NHANES).^3,28^ We also stratified our analysis for two vulnerable populations, men over 50 years of age and postmenopausal women.

## Methods

### Study population

The NHANES is an ongoing survey of the noninstitutionalized U.S. adult population designed to assess their health and nutritional status.^28^ After providing informed consent, participants visited a mobile examination center for standardized physical examination and collection of biological specimens, which were used to assess exposure to environmental chemicals. All study protocols were approved by the National Center for Health Statistics research ethics review board ^3,28^. In our analyses, we included the 2013–2014 NHANES cycle, in which both bone mineral density and serum PFAS concentrations were measured and had not been previously studied. The 2013–2014 cycle also included four (linear and branched) PFAS isomers that were not measured in any previous NHANES cycles. For both evaluations, the selection of NHANES participants was random and designed to maintain the original NHANES characteristics, as previously described.^13,29^ We excluded NHANES participants with missing information about bone mineral measurements (N = 7060) or serum concentrations of PFAS (N = 1450) and those with missing information on covariates (smoking and physical activity) or with bilateral oophorectomy (N = 74). A total of 499 adults (≥40 years) was included in the main analysis. Secondary analyses were performed on 115 men over 50 years and 117 postmenopausal women. Postmenopausal women included women over 60 years old, women who had not had a menstrual period in the previous 12 months. We excluded from the analysis postmenopausal women using hormone replacement treatment or taking parathyroid medication (N=45), due to their influence on the endocrine system (Figure 1).

**Figure 1:**
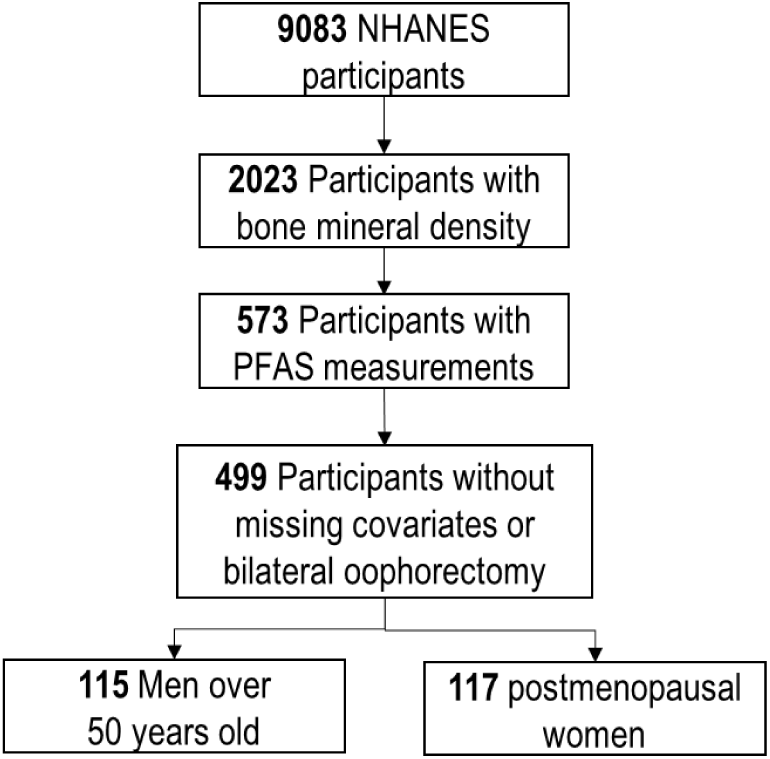
Selection of the National Health and Nutrition Examination Survey (NHANES) participants. Per- and poly-fluoroalkyl substances (PFAS).

### Bone mineral density assessment

Bone mineral density (g/cm^2^) was measured using dual X-ray absorptiometry (Hologic QDR 4500A fan-beam densitometers; Hologic Inc., Bedford, MA, USA).^3^ Antero-posterior lumbar spine mineral density was scanned, and mean density was computed for the first through fourth lumbar vertebrae. For the total and neck femur mineral density, the left hip was routinely scanned. If a left-hip replacement or metal objects in the left leg were reported, the right hip was scanned. Participants were excluded from the femur scan if they had bilateral hip fractures, bilateral hip replacements, or pins. Participants weighing > 300 lbs (136 kg) or pregnant women (defined by self-report or positive urine pregnancy test) were ineligible for the examination. Femur neck has been proposed as the reference skeletal site for defining osteoporosis in epidemiologic studies,^30^ whereas the total femur had been used as a benchmark for osteoporosis in the national Healthy People program ^30^. Each subject’s scan was reviewed in the Department of Radiology, University of California, using standard radiologic techniques and NHANES protocols.^3^

### Serum PFAS measurements

Analysis of 12 PFAS in serum was conducted at the National Center for Environmental Health in a random one-third subsample of nonfasting participants; the NHANES characteristic proportions were maintained.^13,29^ Briefly, serum PFAS were measured using automated solid-phase extraction coupled to isotope-dilution high-performance liquid chromatography– tandem mass spectrometry.^13^ The complete list of PFAS with their acronyms is included in Table S1. In our analyses, we excluded four PFAS because their concentrations were below the limit of detection for more than 60% of samples (Table S1). For the remaining PFAS, when concentrations were less than the limit of detection, a value equal to the limit of detection divided by the square root of two was used in the analyses.

### Statistical methods and analyses

#### The Bayesian WQS (BQWS) regression

Let the values for the correlated mixture components C be scored into quantiles (q_ji_) for the j-th component (j = 1,..,C) and the i-th (i = 1,…,N) participant. We modeled the association between the overall mixture and the outcome y_i_ using a generalized linear model framework

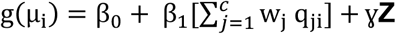

where µ_i_ = E(y_i_), β_0_ is the intercept, and β_1_ is the coefficient mapped to the weighted index 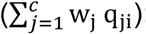; g(.) is link function; and **γ** is a vector of K coefficients mapped to the matrix of K covariates **Z**. Similar to WQS regression, we modeled the weight w_j_ for exposure to the j-th component as an arbitrary function, taking values between 0 and 1, and the sum of all mixture weights to be equal to 1.

The BWQS required specification of the link function, similar to the frequentist approach, and prior probability distributions on all parameters, which differs from WQS regression. The general assumption for each parameter was a weak or uninformative prior. However, more informative priors can be chosen when prior information is known. The link function assumed the following forms:

a. logit link function: y_i_ ∼ binomial (1, µ_i_), when Y was binary;
b. identity link function: y_i_∼Normal(µ_i_, σ^2^), with a noninformative prior for σ^2^∼IGamma(0.1,0.1), when Y was continuous.

Priors for the coefficients were uninformative or weakly informative and were summarized by normal distributions with large variance [β_0_;β_1_∼ Normal(0, 100); γ_k_∼Normal(0, 100)] for each k = 1,…,K. Priors for the weights were modelled as a unit-simplex in order to have non-negative weights (w_j_ Є (0,1)) and to sum all weights to one (Σw_j_ = 1). The natural choice was the Dirichlet distribution with the same density on each vertex for each component of the mixture; i.e., **w** = (w_1_, …, w_C_) ∼ Dirichlet(**α** = (α_1_,…,α_C_)). The **α** parameter can be selected a priori equal to **1**, when investigating the entire domain of the distribution uniformly. Results on simulated datasets, showing the accuracy of the estimates of BWQS, are included in the supplemental material (Figures S3-S5).

#### Statistical Analysis

We estimated the correlations among PFAS and then performed BWQS regression in which bone mineral density (continuous)—in lumbar spine, total femur, and femur neck—was associated with mixtures of PFAS. All analyses were adjusted for race/ethnicity (white, black, Hispanic and other races), age (continuous), sex, physical activity (low-moderate or high), poverty-income ratio (continuous), and smoking status (never or ever smoked). All variables were selected based on previous association with the outcome.^6,7,22^ Priors for all coefficients were uninformative: Normal(0,100). Secondary analyses focused on two vulnerable populations—men over 50 years old and postmenopausal women.

As sensitivity analyses, we assessed the associations between the PFAS mixture and mineral density in lumbar spine, total femur, and femur neck by using the frequentist WQS regression, for the overall population. We also used Bayesian linear regression to determine the association of each PFAS compound with bone mineral density in the overall population. We used R version 3.5.1 for all analyses. All analyses were weighted according to the NHANES weights, appropriately rescaled for the selected subsample, as described on the Centers for Disease Control and Prevention website and in previous studies.^31,32^

## Results

### Study Population Characteristics

Adults included in our main analysis were 55 years old on average (standard error (SE): 0.6), mostly Caucasian (49%), never smoked (60%), reported physical activity below the optimal level (66%), and had an average poverty-income ratio of 2.9 (standard error (SE): 0.9) (Table 1). On average, spine and (total and neck) femur mineral densities were 1.01 (SE: 0.01), 0.95 (SE:0.01), and 0.78 (SE:0.01) g/cm^2^, respectively, in the adult population. Bone mineral density was different by sex, with postmenopausal women having lower density (p<0.05). In all groups, serum concentrations of the linear and branched PFAS isomers (NPFOS, NPFOA, and MPFOS) were higher than all other PFAS levels (PFHS, PFNA, PFDE, MPAH, and PFUA) (Table 1). All serum PFAS concentrations were positively correlated with each other and showed similar patterns across populations (Figure 2).

**Table 1.** NHANES characteristics for the overall adult population, men over 50 years old, and postmenopausal women*

| Characteristics | Overall adult population<br>n=499<br>N = 64,978,899<br>Mean ± SE | Men over 50 years old<br>n=115<br>N = 11,904,882<br>Mean ± SE | Postmenopausal women<br>n=117<br>N = 14,570,823<br>Mean ± SE |
| --- | --- | --- | --- |
| Spine bone density (g/cm<sup>2</sup>) | 1.012 ± 0.009 | 1.047 ± 0.018 | 0.945 ± 0.019 |
| Femur bone density (g/cm<sup>2</sup>) | 0.945 ± 0.007 | 0.989 ± 0.142 | 0.868 ± 0.013 |
| Femur neck density (g/cm<sup>2</sup>) | 0.780 ± 0.007 | 0.794 ± 0.015 | 0.712 ± 0.013 |
| Poverty-income ratio | 2.882 ± 0.090 | 2.893 ± 0.217 | 2.753 ± 0.178 |
| Age (years) | 54.732 ± 0.632 | 62.656 ± 1.022 | 59.410 ± 1.218 |
| Sex: N (%) |  |  |  |
| Male | 24,756,638 (38%) | 11,904,882 (100%) | 0 (0%) |
| Female | 40,222,261 (62%) | 0 (0%) | 14,570,823 (100%) |
| Race: N (%) |  |  |  |
| Mexican American | 8,171,809 (12%) | 1,470,648 (12%) | 2,015,869 (14%) |
| Other Hispanic | 5,506,180 (9%) | 502,741 (4%) | 1,058,314 (7%) |
| Non-Hispanic White | 31,720,096 (49%) | 6,007,804 (51%) | 7,206,459 (49%) |
| Non-Hispanic Black | 11,388,109 (17%) | 2,721,619 (23%) | 2,718,160 (19%) |
| Other | 8,192,706 (13%) | 1,202,068 (10%) | 1,572,021 (11%) |
| Smoking status: N (%) |  |  |  |
| Ever | 25,731,955 (40%) | 6,396,599 (54%) | 4,316,813 (30%) |
| Never | 39,246,944 (60%) | 5,508,283 (46%) | 10,254,011 (70%) |
| Physical activity: N (%) |  |  |  |
| Vigorous | 21,844,374 (34%) | 4,434,527 (37%) | 2,907,339 (20%) |
| Not vigorous | 43,134,525 (66%) | 7,470,355 (63%) | 11,663,484 (80%) |
| PFAS (ug/L): 50<sup>th</sup> (25<sup>th</sup>, 75<sup>th</sup>) percentile |  |  |  |
| NPFOA | 2.00 (1.30, 3.10) | 2.60 (1.90, 3.70) | 1.83 (1.20, 3.00) |
| NPFOS | 4.10 (2.50, 7.40) | 6.21 (3.50, 9.98) | 4.10 (2.70, 7.60) |
| MPFOS | 1.80 (0.90, 3.00) | 3.24 (2.10, 4.30) | 1.70 (1.07, 2.94) |
| MPAH | 0.07 (0.07, 0.20) | 0.10 (0.07, 0.30) | 0.07 (0.07, 0.20) |
| PFDE | 0.20 (0.10, 0.40) | 0.20 (0.10, 0.40) | 0.20 (0.10, 0.34) |
| PFHS | 1.40 (0.80, 2.40) | 2.10 (1.60, 3.56) | 1.30 (0.80, 2.40) |
| PFNA | 0.80 (0.50, 1.30) | 0.90 (0.60, 1.30) | 0.70 (0.50, 1.20) |
| PFUA | 0.10 (0.07, 0.20) | 0.10 (0.07, 0.20) | 0.07 (0.07, 0.20) |

**Figure 2.**
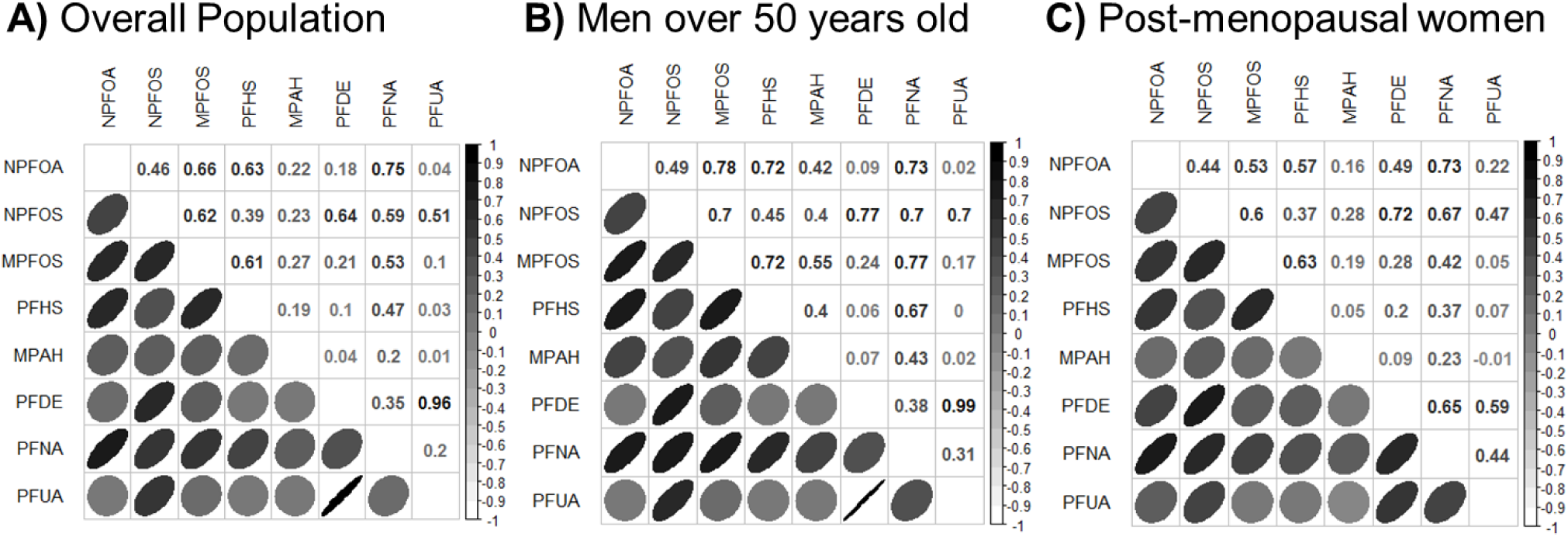
Correlation between serum perfluorinated compounds (PFAS) across the overall adult population, in men over 50 years old, and in postmenopausal women. Color and shape of each elipse reflect the correlation between two compounds.

### BWQS regression characteristics

We employed BWQS regression to identify the association between the mixture of PFAS in subjects’ serum and bone mineral density in spine, total and neck femur, in all adults together and in the most vulnerable populations separately.

PFAS concentrations were categorized into quartiles, and we set the main parameters of the Hamiltonian Monte Carlo chain to optimize the accuracy and speed of the models. In total, we set 1000 iterations, of which 500 were burn-in and 3 were thinned. All parameters showed no autocorrelation between subsequent iterations, and the potential scale-reduction statistics, which were approximately equal to 1 for all estimated parameters, showed convergence of the chain to the equilibrium distributions (Table S2).

### Results in the overall adult population

In the overall adult population there was no evidence of association between the PFAS mixture and bone mineral density in lumbar spine (β=-0.004, 95% credible interval [CrI]: −0.04, 0.04) g/cm^2^, total femur (β =0.002, 95% CrI: −0.04, 0.05) g/cm^2^, or femur neck (β=0.005, 95% CrI: − 0.03, 0.04) g/cm^2^. Components of the mixture contributed approximately equally to the mixture in the outcomes for the overall population (Figures 3A, 4A, and 5A; Table S2).

**Figure 3.**
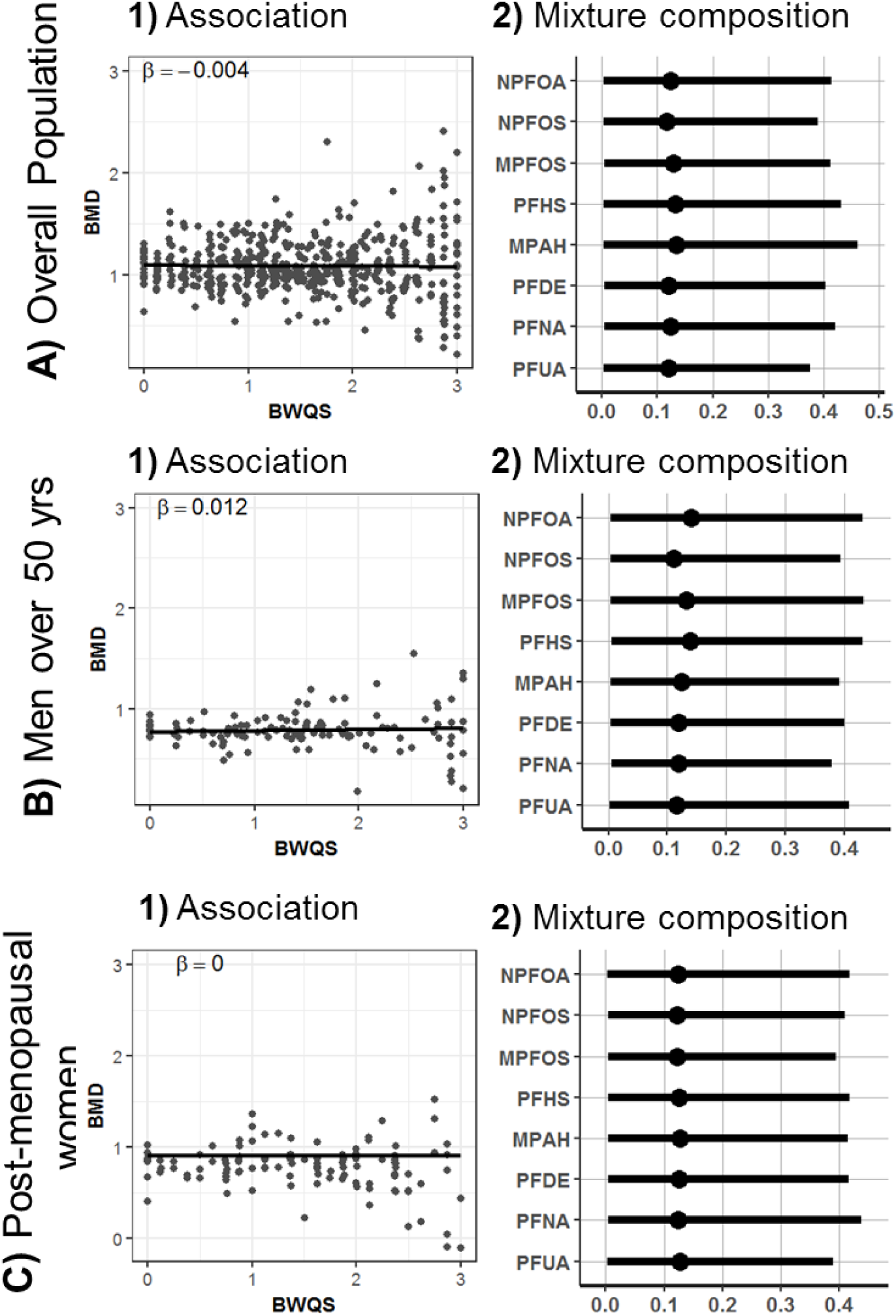
Estimates of the **1)** association between lumbar spine mineral density and the perfluorinated compound (PFAS) mixture and estimates of **2)** mixture composition: weights (percentage rescaled between 0 to 1) with 95% credible intervals for each mixture component in A) the overall population, B) men over 50 years old, and C) postmenopausal women. BMD = Lumbar spine bone mineral density adjusted for race/ethnicity, age, sex, physical activity, poverty-income ratio, and smoking status.

**Figure 4.**
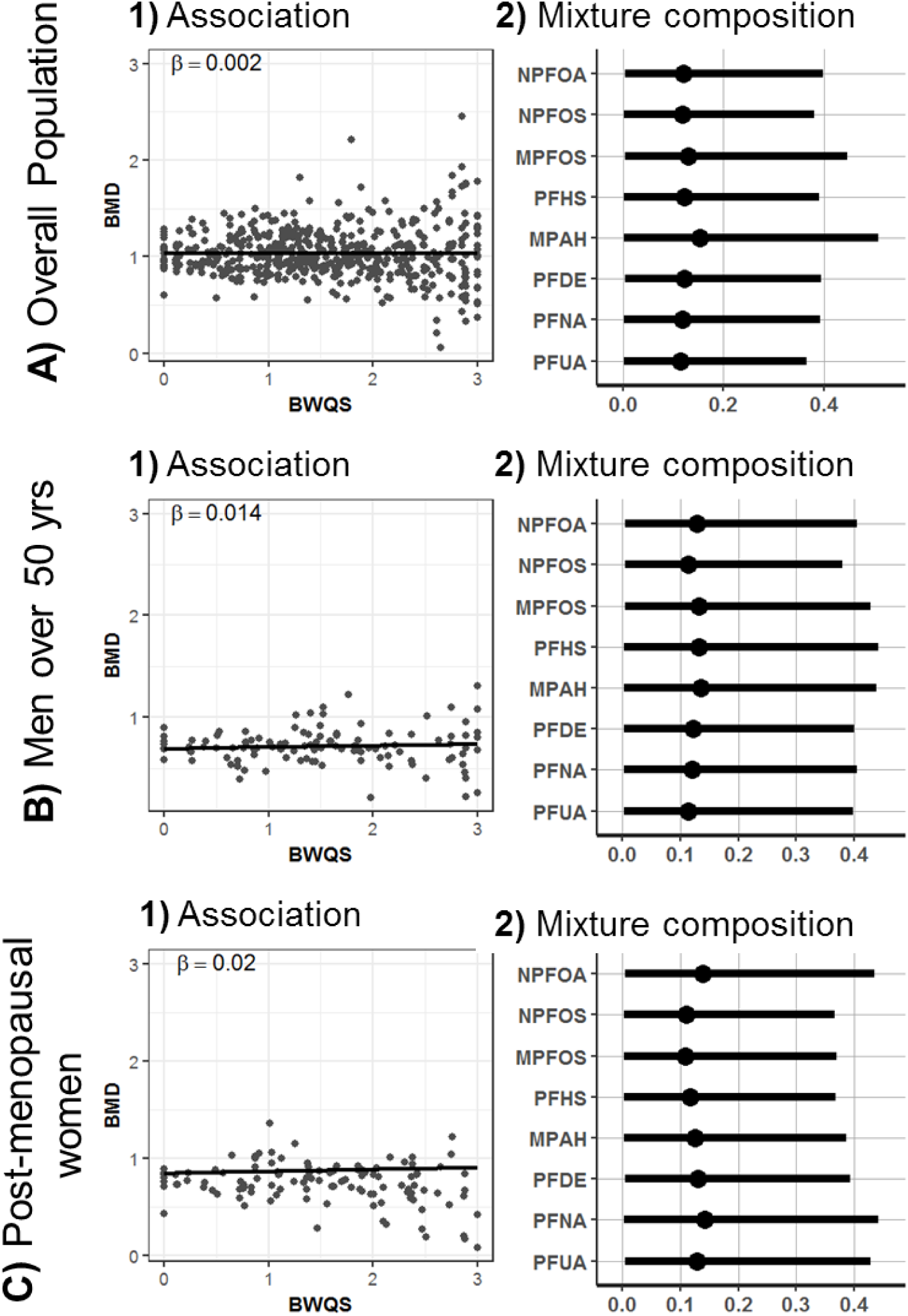
Estimates of the **1)** association between lumbar spine mineral density and the perfluorinated compound (PFAS) mixture and estimates of **2)** mixture composition: weights (percentage rescaled between 0 to 1) with 95% credible intervals for each mixture component in A) the overall population, B) men over 50 years old, and C) postmenopausal women. BMD = total femur bone mineral density adjusted for race/ethnicity, age, sex, physical activity, poverty-income ratio, and smoking status.

**Figure 5.**
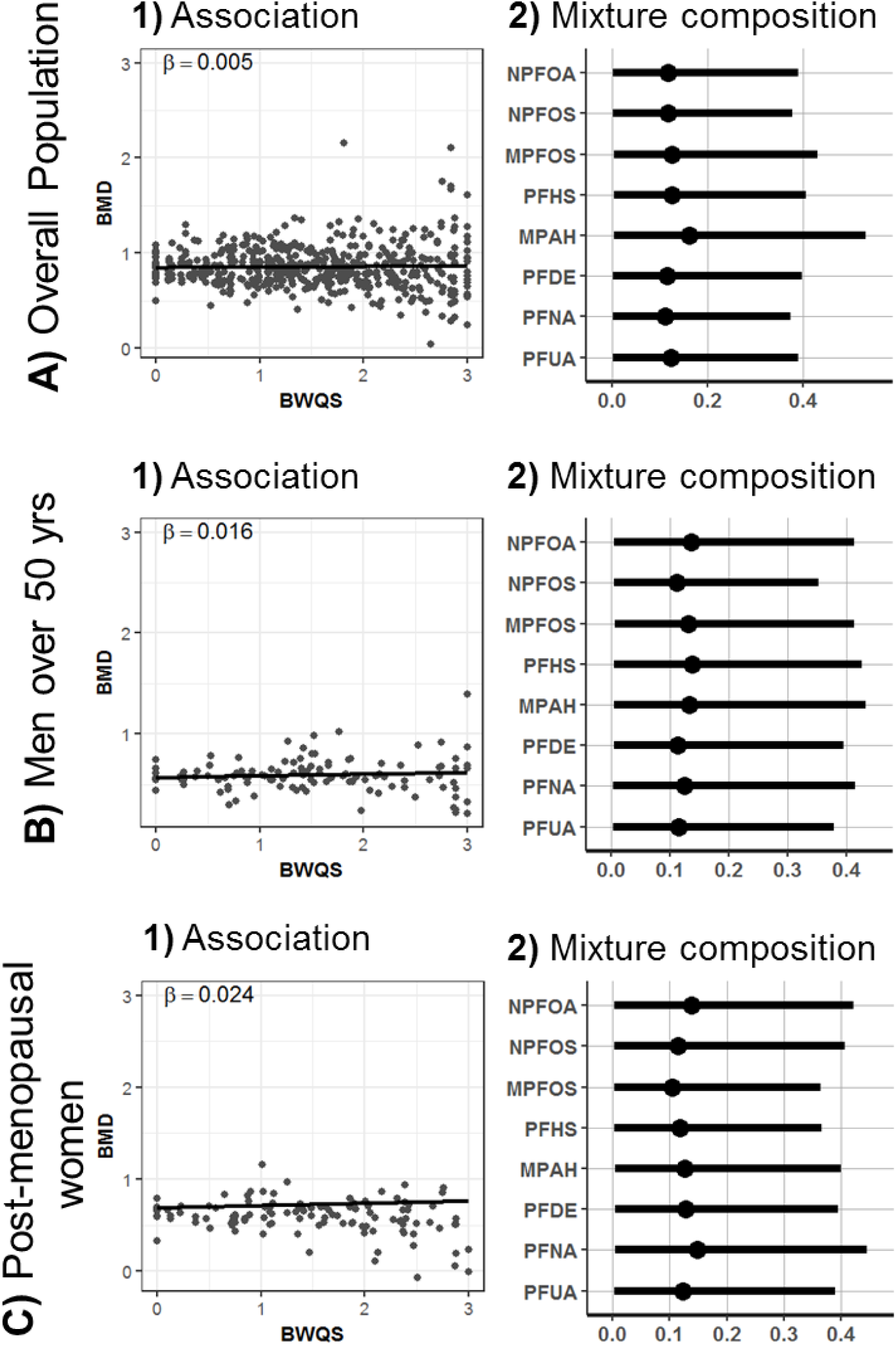
Estimates of the **1)** association between total femur mineral density and the perfluorinated compound (PFAS) mixture and estimates of **2)** the mixture composition: weights (percentage rescaled between 0 to 1) with 95% credible intervals for each mixture component in A) the overall population, B) men over 50 years old, and C) postmenopausal women. BMD = femur neck bone mineral density adjusted for race/ethnicity, age, sex, physical activity, poverty-income ratio, and smoking status.

### Sensitivity analyses in the overall adult population

Results from sensitivity analyses using the frequentist WQS approach, assuming a negative direction between PFAS mixture and bone mineral density, were similar to those of BWQS. We found no associations between the PFAS mixture and all outcomes (lumbar spine: β = −0.01, p = 0.69; total femur: β = −0.01, p = 0.51; femur neck β = −0.004, p = 0.82) using the validation hold out set of 70% (Figure S1, Table S3). Individual PFAS analyses showed a weak negative association among MPFOS and all bones (lumbar spine: −0.02, 95% CrI: −0.03, −0.009; total femur: −0.01, 95% CrI: −0.02, −0.003; and femur neck −0.01, 95% CrI: −0.02, −0.004), whereas PFNA was positively associated with total femur (0.04, 95% CrI: 0.01, 0.06) and neck femur (0.03, 95% CrI: 0.01, 0.05) (Figure S2, Table S4). However, those associations did not persist after correcting for multiple comparisons (data not shown).

### Results in men over 50 years old and in postmenopausal women

There was no evidence of an association between the PFAS mixture and bone mineral densities in men over 50 years old (lumbar spine: β =0.01, 95% CrI: −0.05; 0.08; total femur β =0.01, 95% CrI: −0.04, 0.07; and femur neck β =0.02, 95% CrI: −0.04, 0.07) or in postmenopausal women (lumbar spine β =0.00, 95% CrI: −0.08, 0.08; total femur β =0.02, 95% CrI: −0.04, 0.08; and femur neck β =0.02, 95% CrI: −0.03, 0.08). The contribution of all mixture components was similar across bones and across populations (Figures 3-5 panels B and C; Table S2).

## Discussion

We extended the WQS regression under a Bayesian framework and determined the combined association of eight PFAS with bone mineral density in lumbar spine and total and neck femur in a survey representative U.S. adult population in the years 2013–2014. The main BWQS characteristics include diagnostic statistics for all estimated parameters, and BWQS does not require a priori selection of the directionality of the coefficient associated with the mixture or splitting the original dataset, thus overcoming a few limitations of the frequentist approach.

The application of our novel method showed no evidence of the association between serum concentration of PFAS mixture, composed of linear and branched PFOS and PFOA isomers (NPFOS, NPFOA, and MPFOS) and PFHS, PFNA, PFDE, MPAH, and PFUA, and bone mineral density in lumbar spine and total and neck femur. The contribution of each compound was similar across bones. Results were also consistent using the frequentist WQS approach and Bayesian linear regressions.

Our results confirmed prior findings showing null associations between PFAS exposure and bone health in the overall adult U.S. population. Serum concentrations of individual PFAS, namely, PFOA, PFOS, PFHS, and PFNA, were not associated with mineral density in spine and total and neck femur in the overall adult population in the 2009–2010 NHANES cycle.^22^ Previous findings showed that associations between a few individual PFAS compounds and bone mineral density were negative and significant in women only, with higher concentrations of PFOA, PFOS, PFHS, and PFNA associated with lower bone mineral density and with a higher risk osteoporosis in women.^22^ However, those analyses ignored the correlations among those compounds and did not consider PFAS as a mixture, thereby increasing the number of false positives among significant results.

Epidemiological studies support the hypothesis that PFAS are endocrine disruptors. PFAS modulate thyroid and sex hormone concentrations, which play a critical role in bone remodeling and health ^33^. Chronic PFAS exposure was associated with suppressed serum thyroxine and triiodothyronine (T3) levels in human studies ^34,35^ and with altered responses to T3 in a T3-dependent cell line *in vitro*.^36^ A cross-sectional study of adult NHANES participants ^35^ reported that serum PFHS was positively associated with subclinical hyperthyroidism in women, which is a risk factor for decreased bone mass.^37^ PFAS also interfere directly with estrogen and androgen receptors, disrupting the biological effects of sex hormones and leading to reduced fecundity in women and delayed puberty in boys, both of which are associated with lower bone mineral density later in life.^23,38^ In studies of women’s health, PFAS were associated with decreased production of estradiol and progesterone, which are essential hormones for bone health due to their promotion of osteoblast activity. Based on this body of literature, the PFAS mixture, mediated by endocrine receptors, might affect bone mineral density, but further studies with longitudinal design are required to address this problem.

We also found no evidence of an association between the PFAS mixture and bone mineral density in vulnerable populations, including men over 50 years old and postmenopausal women. However, our results could have been limited by a small number of participants (n = 115 and 117, respectively). Also, men and women in this study were not asked whether they attempted to prevent bone deterioration by changes in lifestyle, such as using supplements or alternative treatments or eating a healthier diet.

PFOA and PFOS, both of which were previously associated with bone mineral density in women ^19,22^ were analyzed and decomposed in linear and branched isomers in our analyses, and we could not confirm previous results for those compounds. Due to the cross-sectional design of our study, we could not rule out whether it was reverse causation and PFAS exposure preceded the outcomes or bone mineral status affected the response to exposure ^39^. It is also possible that our results may have been limited by the smaller number of participants (n=499) than other study showing an association between the exposure of a few PFAS and bone mineral density (n=1914).^22^ Further studies with longitudinal design and larger sample size could help to disentangle the underlying mechanism linking PFAS exposure and bone health in older adults.

The strengths of our study include a novel statistical approach, which accounted for the correlation among co-occurring PFAS and provided information about the overall adverse associations of PFAS and bone density. We used a sample that is known to represent the U.S. population in the years 2013–2014, and we relied on PFAS concentrations and bone mineral density levels that were validated and compared across NHANES cycles.

## Conclusions

This is the first study to assess the relationship between exposure to a mixture of PFAS and bone mineral density at three bone sites. The novel Bayesian WQS approach identified both the overall association between PFAS mixture with bone mineral density and the contribution of each PFAS to the mixture. The serum PFAS mixture showed no association with mineral density of the lumbar spine and total and neck femur in the NHANES adult population in the years 2013–2014.

## Data Availability

Data are available on the Centers for Disease Control and Prevention (CDC)’s website (National Health and Nutrition Examination Survey cycle 2013-2014).

## Financial support and acknowledgments

This research was supported by grant P30ES023515, U2CES026444, UH3OD023337-03 of the National Institute of Environmental Health Sciences of the National Institutes of Health. The authors would like to thank Dr. Christine Schubert Kabban for helpful comments.

